# Prefrontal tDCS for improving mental health and cognitive deficits in patients with Multiple Sclerosis: a randomized, double-blind, parallel-group study

**DOI:** 10.1101/2024.05.19.24306880

**Authors:** Nasim Zakibakhsh, Sajjad Basharpoor, Hamidreza Ghalyanchi Langroodi, Mohammad Narimani, Michael A Nitsche, Mohammad Ali Salehinejad

## Abstract

**Background:** Multiple Sclerosis (MS) is an autoimmune disease associated with physical disability, psychological impairment, and cognitive dysfunctions. Consequently, the disease burden is substantial, and treatment choices are limited. In this randomized, double-blind study, we used repeated prefrontal electrical stimulation and assessed mental health-related variables (including quality of life, sleep, psychological distress) and cognitive dysfunctions (psychomotor speed, working memory, attention/vigilance) in 40 patients with MS.

**Methods:** The patients were randomly assigned (block randomization method) to two groups of sham (*n*=20), or 1.5-mA (*n*=20) transcranial direct current stimulation (tDCS) targeting the left dorsolateral prefrontal cortex (F3) and right frontopolar cortex (Fp2) with anodal and cathodal stimulation respectively (electrode size: 25 cm^2^). The treatment included 10 sessions of 20 minutes stimulation delivered every other day. Outcome measures were quality of life, sleep quality, psychological distress, and performance on a neuropsychological test battery dedicated to cognitive dysfunctions in MS (psychomotor speed, working memory, and attention). All outcome measures were examined pre-intervention and post-intervention. Both patients and technicians delivering the stimulation were unaware of the study hypotheses and the type of stimulation being used.

**Results:** The active protocol significantly improved quality of life and reduced sleep difficulties and psychological distress compared to the sham group. The active protocol, furthermore, improved psychomotor speed, attention and vigilance, and some aspects of working memory performance compared to the sham protocol. Improvement in mental health outcome measures was significantly associated with better cognitive performance.

**Conclusions:** Modulation of prefrontal regions with tDCS ameliorates secondary clinical symptoms and results in beneficial cognitive effects in patients with MS. These results support applying prefrontal tDCS in larger trials for improving mental health and cognitive dysfunctions in MS.

**Trial registration:** ClinicalTrials.gov Identifier: NCT06401928

## 1. Introduction

Multiple Sclerosis (MS) is the most common autoimmune disorder of the central nervous system, afflicting more than 2.5 million people worldwide, especially young people (1, 2). It is a progressive chronic disease, caused by an autoimmune attack, which results in the gradual loss of the myelin sheath around neuronal axons of the central nervous system. Depending on the affected neurons, different symptoms are expressed in the course of the disease; nonetheless, physical disability, cognitive impairment, and decreased quality of life are common consequences of the disease (3). MS results in motor, sensory, cognitive, and neuropsychiatric symptoms, all of which can occur independently of one another (4). The disease is associated with a high mental health burden due to primary symptoms of sensory and motor deficits. Some of the most common symptoms include fatigue, vision problems, numbness and tingling, muscle spasms, stiffness and weakness, mobility problems, and pain (2, 5).

In addition to primary symptoms such as fatigue, motor deficits, and visual disturbances, people with MS also experience disability in mental health variables, including quality of life, sleep, and emotional disturbances (6–8). Cognitive deficits are also commonly observed symptoms in MS and include deficits in attention and vigilance, information processing, executive functioning, processing speed, and long-term memory (4) with a profound effect on instrumental activities of daily living (9). Both primary and secondary symptoms can vary in severity and may come and go, depending on disease activity, progression, and the type of MS, which include relapsing-remitting, or chronic-progressive dynamics (8, 10). Individuals with MS need to work closely with healthcare professionals to manage their symptoms and optimize their quality of life. Considering the burden of treatment (11), the complex pathophysiology and psychophysiology of MS, and limited treatment options for secondary symptoms (physical therapy, psychotherapy, cognitive rehabilitation) (3), there is a need for novel and effective treatment for both primary and secondary symptoms.

Transcranial direct current stimulation (tDCS) is a safe and easy-to-use noninvasive brain stimulation intervention for studying and modifying human brain functions (12, 13). It involves applying a low-intensity direct current to the scalp, which induces alterations of the resting membrane potential of neurons. At the macroscale level, anodal stimulation depolarizes neurons at a subthreshold level, making them more likely to fire action potentials, while cathodal stimulation hyperpolarizes neurons, reducing their excitability (14, 15). By changing the excitability of the brain and inducing neuroplasticity effects, it is possible to restore functional brain abnormalities and affect target behavior/cognition. Previous studies have shown functional brain abnormalities in MS that are partially related to secondary symptoms in MS (16, 17). These abnormalities can take many different forms, such as modified brain activity patterns, neural network disturbances, and modifications of cognitive functions. Key elements of functional brain impairments in MS include changes in pain processing pathways as well as alterations of brain regions involved in the regulation of mood and arousal, including the limbic system, hypothalamus, and motor regions (18–20). Furthermore, there is growing evidence of grey matter changes in MS patients that are linked to disability and other clinical symptoms in these patients (21, 22).

Previous tDCS studies have mostly focused on examining the efficacy of prefrontal and motor tDCS on clinical symptoms (e.g., fatigue, pain), motor symptoms, and cognitive deficits of patients with MS (23–26). There is inconsistency in study results regarding the cognitive effects of non-invasive brain stimulation, such as tDCS, on MS patients. In addition to cognitive and motor symptoms, MS patients also suffer from other mental health issues that are less commonly addressed in tDCS studies. While there have been some studies on the effects of transcranial direct current stimulation (tDCS) on mental health-related variables, such as quality of life (27), sleep (28), and emotional difficulties (29), there is still a lack of research on the impact of tDCS on both mental health-related variables and cognitive performance in patients with MS. Accordingly, the purpose of this study is to address this research gap by examining 1) the effects of repeated prefrontal tDCS on mental-health-related variables (i.e., quality of life, sleep difficulties, psychological distress) in patients with MS and 2) the effects of repeated prefrontal tDCS on the cognitive performance of patients with MS in a randomized, double-blind, sham-controlled trial.

## 2. Methods

### 2.1. Participants

Eighty MS patients from the local community (Rasht, Iran) were screened for inclusion in the study. The inclusion criteria were: (1) diagnosis of MS according to the diagnostic criteria for multiple sclerosis: 2010 Revisions of the McDonald criteria (Polman et al., 2011), certified by a professional neurologist, (2) being 25–55 years old, (3) being non-smoker, (4) no previous history of neurological diseases, brain surgery, epilepsy, seizures, brain damage, head injury, or metal brain implants, (5) absence of other psychiatric disorders except mood and anxiety disorders, and (6) no relapse of MS symptoms within the last two months before beginning the experiment. Of those who met the inclusion criteria (*n*=60), forty patients were randomly assigned to the experimental (active tDCS) and control (sham tDCS) groups based on a sample size analysis (f = 0.30, α = 0.05, power = 0.95, mixed-model ANOVA for 2 groups with 2 measurements) which resulted in a sample size of 40 patients. For the group assignment of the participants, block randomization method was applied. Three patients decided to withdraw from the study following the first session and thus the final analysis was conducted on 37 patients (mean age = 37.30, SD = 6.21, 27 females, 10 males) (see Table 1 and Figure 1 for demographics and study inclusion). This is a retrospectively registered clinical trial (ClinicalTrials.gov Identifier: NCT06401928) approved by the Ethics Committee of the Mohaghegh Ardabili University (Ethics code: IR.MAU.REC.1401.94). Participants gave their written informed consent before participation.

**Figure 1.**
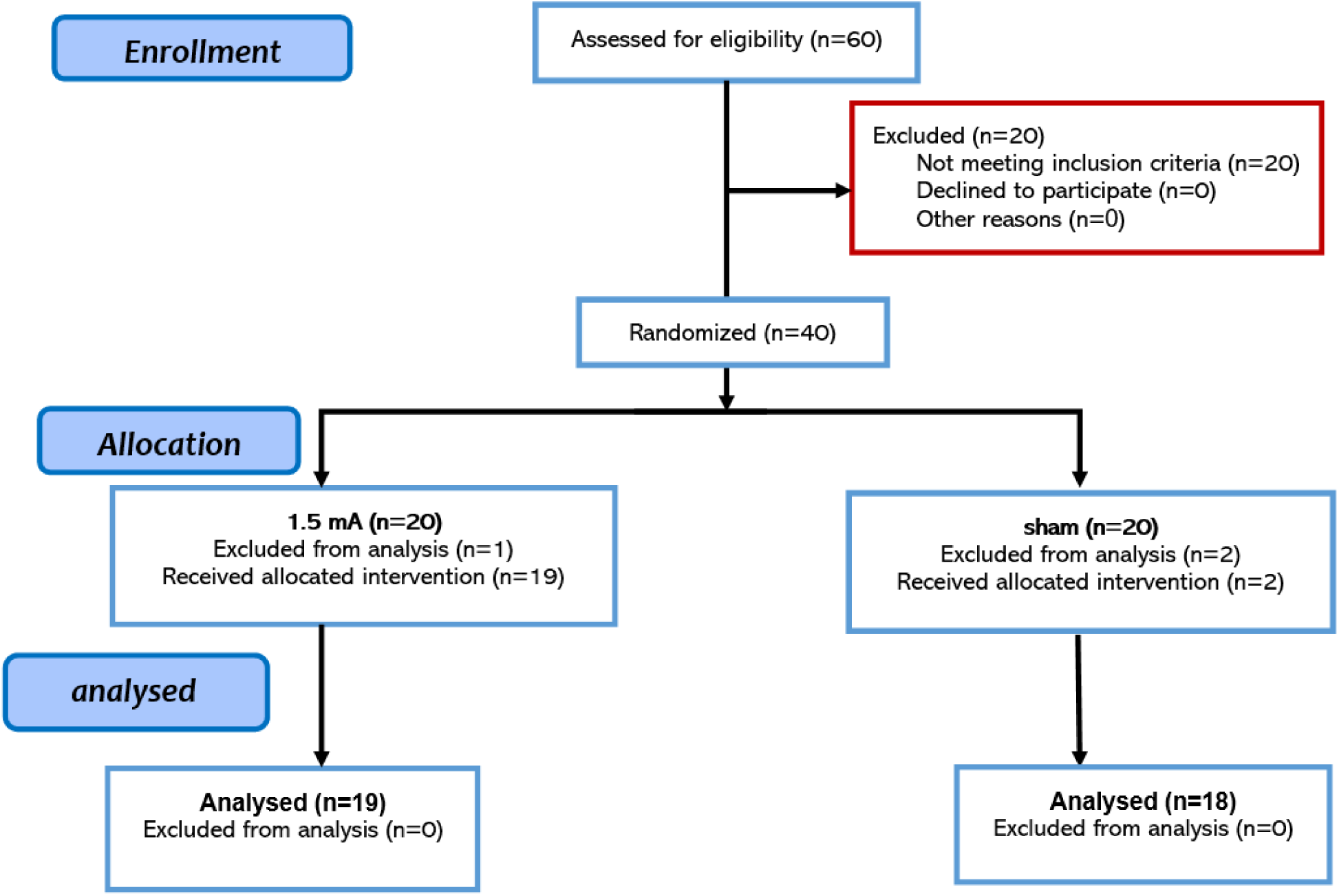
displays the CONSORT Flow Diagram illustrating the study’s inclusion procedures. Thirty-seven patients successfully completed the post-intervention measurement for all CANTAB subtests.

**Table 1.** Demographic data.

|  | Active tDCS | Sham tDCS | *p-value\** |
| --- | --- | --- | --- |
| Sample size (n) | 19 | 18 |  |
| Age- M(SD) | 36.89(1.47) | 37.7(1.45) | 0.691 |
| Sex – Male (female) | 4(15) | 6(12) | 0.401 |
| Marital Status – Single (married) | 9(10) | 7(11) | 0.603 |
| OAID yes (No) | 4(15) | 1(17) | 0.340 |
| Family history- yes (No) | 9(10) | 6(12) | 0.508 |
| **Duration of disease** |  |  | 0.078 |
| 1-10 years | 14 | 7 |  |
| 11-20 years | 4 | 10 |  |
| More than 20 years | 1 | 1 |  |
| **Type of MS** |  |  | 0.208 |
| CIS | 3 | 5 |  |
| RR | 14 | 13 |  |
| SR | 2 | 0 |  |
| **Type of medications** | 10(7) | 12(5) | 0.663 |
| IS (IM) |  |  |  |

*Note:* tDCS = transcranial Direct Current Stimulation; M = Mean; SD = Standard Deviation; OAID = other .autoimmune. disease; CIS= clinically isolated syndrome; RR=relapsing-remitting multiple sclerosis; SP=secondary progressive multiple sclerosis; IS= immunosuppressant drug; IM=immunomodulatory drug; \* = between-group differences in demographic variables were explored by Chi-square tests or Fisher's exact test for categorical variables and F-tests for continuous variables.

### 2.2. Measures

#### 2.2.1. Mental health assessments

The primary outcome measures of the study included quality of life, sleep difficulties, and psychological distress that were measured with the Multiple Sclerosis Impact Scale (MSIS-29) (30), the Mini Sleep Questionnaire (MSQ) (31), and the Depression Anxiety Stress Scale-21 (DASS-21) (32) respectively. The MSIS-29 is a measure of the physical and psychological impact of MS from the patients’ perspective consisting of 29 questions with the first 20 items focusing on the physical impact and the remaining 9 on the psychological impact. The Mini Sleep Questionnaire (MSQ) is typically used to screen sleep disturbances in clinical populations, and the DASS-21 is a 21-item self-report measure designed to assess the severity of general psychological distress and symptoms related to depression, anxiety, and stress in adults and older adolescents (+17 years). Details about these measures and their psychometric properties can be found in the supplementary information. A native-language version of each test was used in this study (33–35).

#### 2.2.2. Neuropsychological performance

In addition to mental health-related outcome variables, we assessed the neuropsychological performance and cognitive functioning of the MS patients with several subtests of a neuropsychological battery designed for MS patients using the Cambridge Cognition Neuropsychological Battery (CANTAB). The battery included 3 computerized tests to measure psychomotor speed, working memory and sustained attention / vigilance via the Reaction Time (RTI), Spatial Working Memory (SWM) and Rapid Visual Information Processing (RVP) tests respectively. The tests included in this battery are highly sensitive to cognitive impairments in MS across the disease severity spectrum (36). Details about these measures are available in the supplementary information.

### 2.3. tDCS

We used a two-channel Neurostim stimulator device (MadinaTeb, Tehran, Iran) powered by a 9-volt alkaline battery. Electrical current was applied through a pair of saline-soaked sponge electrodes (5 × 5 cm) for a period of 20 minutes (with 30 seconds ramping up and 30 seconds ramping down) and a stimulation intensity of 1.5 mA. We had two stimulation conditions: active and sham tDCS. In the active condition, the anodal electrode was placed over the left dorsolateral prefrontal cortex (left DLPFC) over the F3 electrode position according to the 10-20 EEG International System and the cathodal electrode was placed over the right frontopolar cortex (FPC) (electrode position Fp2) which includes orbitofrontal cortex (BA 10, 11) (37, 38). To reduce shunting of current between the electrodes through the scalp, the edges had a distance of at least 6 cm. In the sham condition, a sham stimulation was employed with the same electrode configuration. Here, the electrical current was ramped up for 30 seconds followed by 30 seconds of stimulation and 30 seconds of ramping down to generate the same sensation as the active condition, and then was turned off without the participants’ knowledge. This method of sham stimulation has been proven reliable (39). During the study, participants were blinded to the type of stimulation they received. A survey was conducted after each session to document any reported side effects, but blinding efficacy (asking participants to guess about the type of stimulation) was not explored due to the multi-session design of the study. After finalizing the study protocol, also the patients in the sham group were assigned to active tDCS intervention, but the latter procedure was beyond the focus of the study protocol.

### 2.4. Procedure

Prior to the experiment, participants completed a brief questionnaire to evaluate their suitability for brain stimulation. The active tDCS and sham groups received 10 sessions of stimulation (three sessions per week, resulting in a total of three weeks plus an additional day) with 24-hour between-session intervals (except for the 4th and 8^th^ sessions that had 48-h interval due to the weekend). Clinical measures (psychological distress questionnaires and cognitive assessment) were evaluated before the first tDCS session (pre-intervention), and right after the end of the last tDCS session (post-intervention). All tDCS sessions were scheduled between 2:00 and 5:00 p.m. and were monitored for sleep pressure to mitigate a potential impact of circadian variation on cortical excitability and neuroplasticity induction for all participants across all sessions (40, 41). Before starting the experiment, the participants were given instructions about the cognitive tasks. The measurement sessions before the intervention and after the intervention took about 2 hours. Each session was conducted on two separate days to prevent fatigue. The first day was dedicated to clinical assessment (quality of life, sleep, psychological distress), and the second day for cognitive assessment (psychomotor speed, working memory, attention/vigilance). To ensure that the experiment was double-blinded, independent researchers who were not involved in the administration of stimulation sessions were responsible for supervising the examination of outcome measures, data analysis, and group assignment (see Fig. 2).

**Figure 2:**
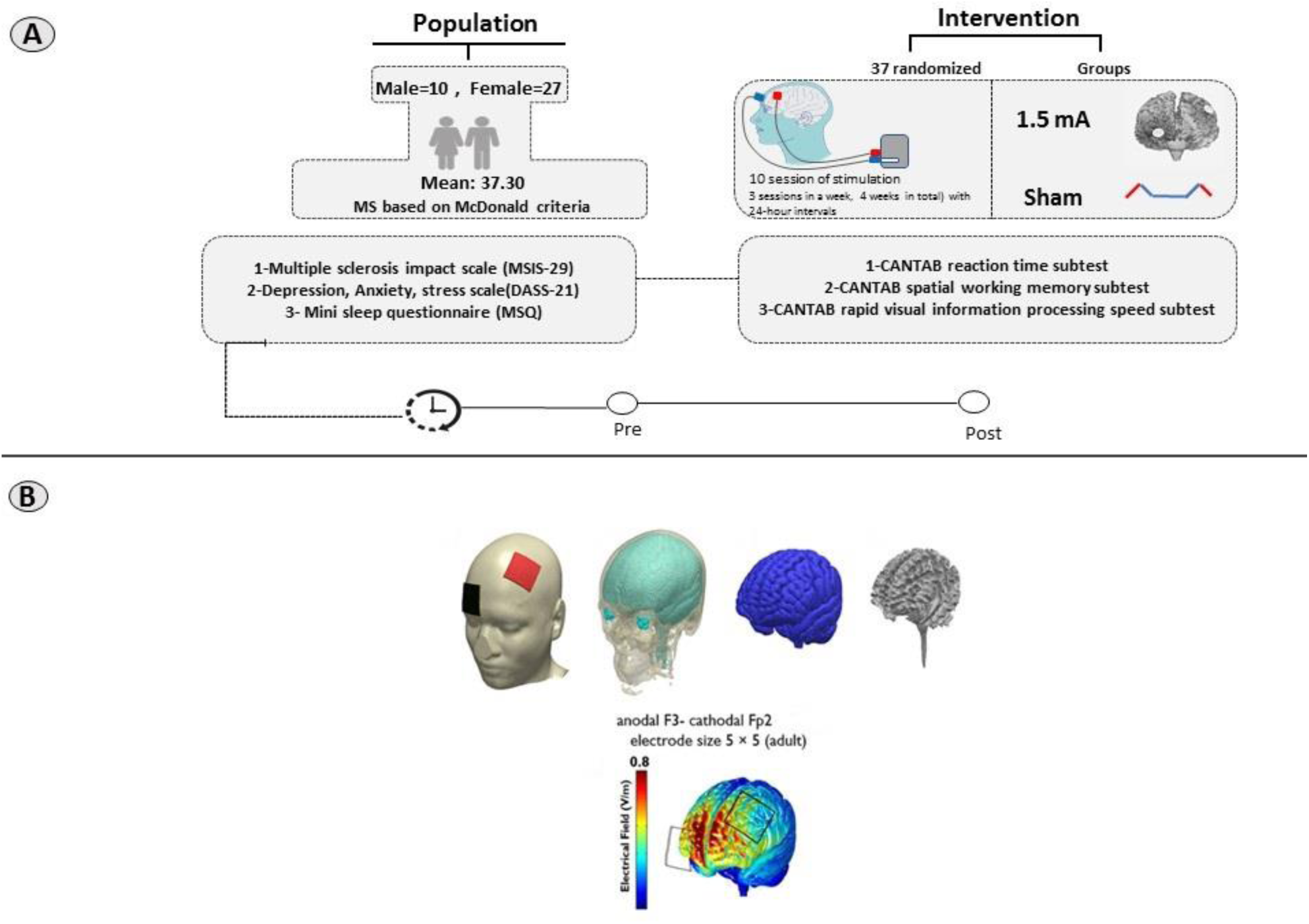
Study Design: (A) This study was conducted as a randomized, double-blind trial. Participants were divided into two groups: active tDCS (n = 19) and sham tDCS (n = 18). Both groups underwent pre- and post-intervention assessments. The anodal electrode was positioned over the left dorsolateral prefrontal cortex (DLPFC), while the cathode was placed over the right frontopolar cortex (FPC). (B) 3D models were utilized to examine the flow of electrical current in the brain following the specified protocol. The MR image was segmented into six tissue types: gray matter (GM), white matter (WM), CSF, skull, scalp, and air cavities using SPM8 from the Welcome Trust Center for Neuroimaging, London, UK, with an enhanced tissue probability map. The segmented images were used to create a 3D model with Simpleware software version 5 from Synopsys, Mountain View, CA, incorporating the electrodes and saline-soaked sponges. The distribution of current flow within the brain was then computed using the finite element method in COMSOL Multiphysics software version 5.2 from COMSOL Inc., Burlington, MA. The electric fields were visualized for stimulation intensities of 2.0 mA with an F3 anodal–Fp2 cathodal montage. Please note: This model illustrates the current flow for 2 mA tDCS for illustrative purposes. The induced electric field of 1.5 mA differs from the 2 mA field.

### 2.5. Statistical analysis

Data were analyzed with the statistical package SPSS, version 27.0 (IBM, SPSS, Inc., Chicago, IL). The normality and homogeneity of data variance were confirmed by Shapiro-Wilk and Levin tests, respectively. Mixed model ANOVAs were conducted for the dependent variables (MSIS, DASS, MSQ scores; psychomotor speed, working memory and attention tasks) with “group” (active vs sham) as the between-subject and time (pre-intervention, post-intervention) as the within-subject factors. Mauchly’s test was used to evaluate sphericity of the data before performing the respective ANOVAs (*p*<0.05). In case of violation, degrees of freedom were corrected using Greenhouse-Geisser estimates of sphericity. Post hoc comparisons were conducted with Fisher’s LSD post-hoc tests for individual mean difference comparisons across groups (active 1.5 mA, sham) and time points (pre-intervention, post-intervention). Pearson’s correlation was also calculated to explore potential associations between mental health-related variables and cognitive performance. The critical level of significance was 0.05 for all statistical analyses.

## 3. Results

### 3.1. Data overview

Demographic information is summarized in Table 1. Patients tolerated the stimulation well and no adverse effects were reported during and after stimulation. No significant difference was found between the group ratings of tDCS side effects except for the burning sensation (*p*=0.044) which was higher in the active group (Table 2). The burning sensation, however, did not correlate with any outcome measure, thus ruling out respective effects. No significant between-group differences were observed for baseline measurements of outcome variables (Table 3).

**Table 2:** Means and SDs of reported tDCS side effects.

|  |  | active tDCS | Sham | t | df | p-value* |
| --- | --- | --- | --- | --- | --- | --- |
|  |  | M(SD) | M(SD) |  |  |  |
| **Reported side effect** | Skin redness | 0.32(1.003) | 0.11(0.32) | 0.844 | 21.888 | 0.408 |
|  | sleep | 0.32(1.15) | 0.0(0.0) | 1.189 | 18 | 0.25 |
|  | fatigue | 0.26(1.14) | 0.11(0.47) | 0.532 | 24.176 | 0.599 |
|  | pain | 0.21(0.53) | 0.17(0.38) | 0.288 | 32.642 | 0.775 |
|  | burning | 1.32(1.63) | 0.44(.705) | 2.124 | 24.747 | **0.044** |
|  | itching | 0.84(1.25) | 0.56(0.85) | 0.813 | 31.822 | 0.422 |
|  | tingling | 1.05(1.35) | 1(0.907) | 0.14 | 31.608 | 0.89 |

Values are presented as means ± standard deviation (SD). Note: each value represents the average of side effects reported during all 10 tDCS sessions. tDCS = transcranial Direct Current Stimulation; M = Mean; SD = Standard Deviation; \* = between-group differences of side effect items were explored by independent samples t-tests, Significant results are highlighted ( $p \leq 0.05$ ) in **bold**

**Table 3:**
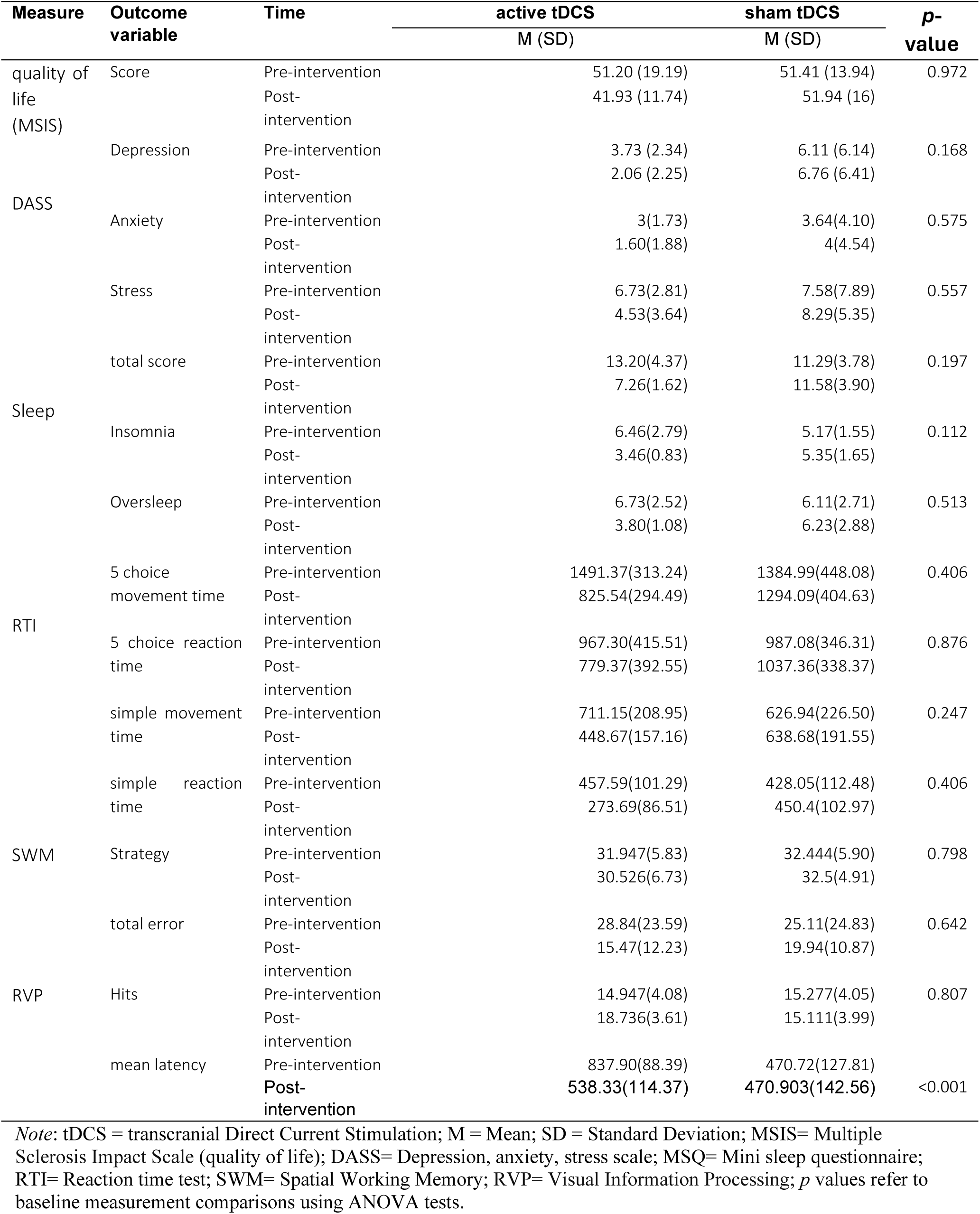
Means and SDs of outcome variables before and after the tDCS.

### 3.2. Efficacy of tDCS on quality of life, sleep quality, and psychological distress

The results of the 2 (group) × 2 (time: pre, post) mixed model ANOVA showed a significant main effect of time (*F_1_*=7.94, *p*=0.008, *η*p2=0.21) and a significant group×time interaction (*F_1_*=9.98, *p*=0.004, *η*p2=0.25) but no main effect of group (*F_1_*=0.94, *p*=0.338) on quality of life scores measured by the MSIS-29. Fisher’s LSD post-hoc tests showed a significant increase in quality of life scores in patients with MS who received active tDCS after the intervention compared to pre-intervention (*t*=4.74, *p*<0.001). When compared to the sham group, active tDCS significantly improved *quality of life* scores after the intervention (*t* =4,93, *p*<0.001). Baseline between-group comparisons (active groups vs sham) showed no significant differences in the pre-intervention scores (Table 4, Fig. 3A, B).

**Figure 3:**
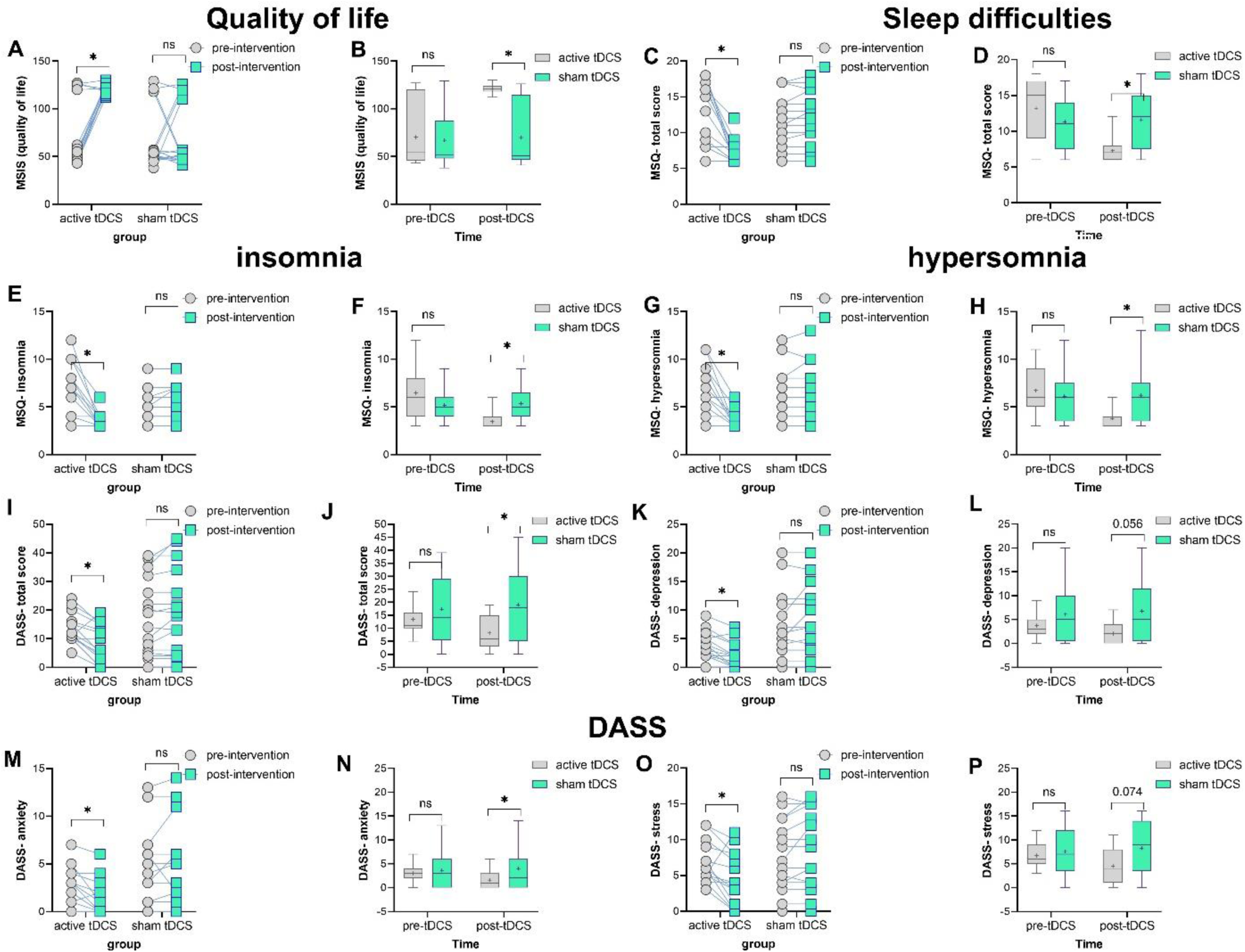
The effect of prefrontal active and sham tDCS on mental health-related variables. The left panel displays average outcome measures before and after the intervention within each group (i.e., within-group comparisons). The right panels illustrate average outcome measures for each time point (i.e., pre-intervention or post-intervention) across groups (active vs sham). *Note*: tDCS = transcranial Direct Current Stimulation; DASS = Depression Anxiety Stress Scale; Floating asterisks [*] in the left panel represent a significant difference between pre-intervention measurements vs post-intervention measurements in all groups. Floating asterisks [*] in the right panel indicate a significant difference between active stimulation (1.5 mA) vs sham tDCS at each time point. ns non-significant. All pairwise comparisons were conducted using Fisher’s LSD multiple comparisons tests. All error bars represent s.e.m.

**Table 4:**
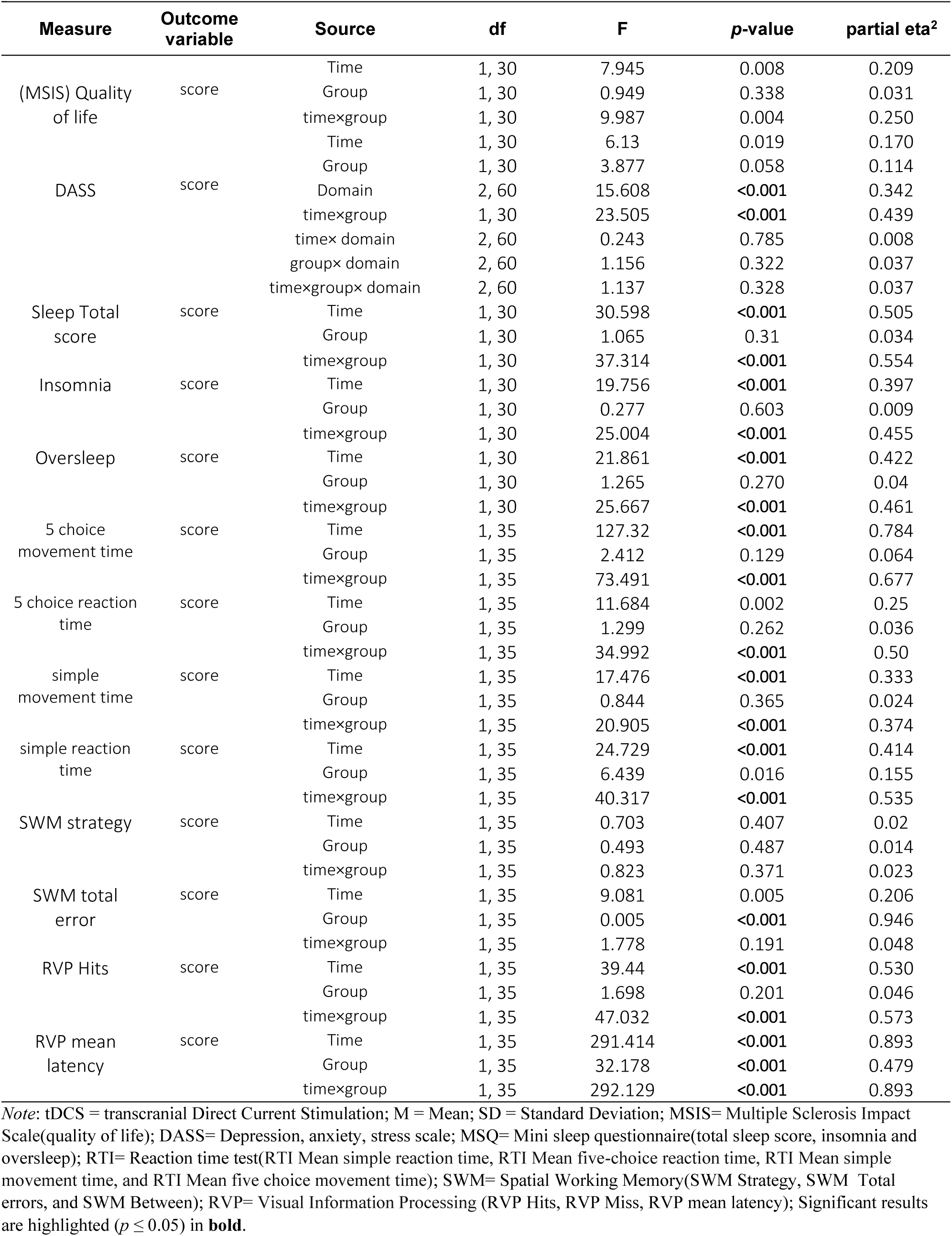
Results of the mixed model ANOVAs for effects of group (active tDCS, sham) and time (pre-intervention, post-intervention) on outcome measures.

With respect to the sleep quality scores measured by the MSQ, the results of a 2 × 2 mixed model ANOVA showed a significant main effect of time (*F_1,30_*=30.598, *p*<0.001, *η*p2=0.505) and a significant group×time interaction (*F_1,30_*=37.314, *p*<0.001, *η*p2=0.554) but no main effect of group (*F_1,30_*=1.065, *p*=0.31). Fisher’s LSD post-hoc tests showed a significant improvement of sleep quality scores only in patients with MS that received active tDCS after the intervention compared to pre-intervention (*t_total sleep_* =4.50, *p*<0.001, *t_insomnia_* =4.48, *p*<0.001, *t_hypersomnia_* =3.29, *p*<0.001) but not in the sham group. When compared to the sham group, the active tDCS group significantly improved with respect to the sleep total score (*t*=3.38, *p*=0.001), insomnia (*t*=2.90, *p*=0.005) and hypersomnia (*t*=2.82, *p*=0.006). Baseline between-group comparisons were not significant (*p*>0.05) (Fig. 3 C-H)

For psychological distress, a 3 (domain: depression, anxiety, stress) × 2 (time: pre, post) × 2 (group) mixed model ANOVA was conducted. The results showed significant main effects of time (*F_1,30_*=6.13, *p*=0.019, *η*p2=0.17), domain (*F_2,60_*=15.608, *p*<0.001, *η*p2=0.342) and a significant group×time interaction (*F_1,30_*=23.505, *p*<0.001, *η*p2=0.439). The main effect of group, and the domain×time, domain×group and domain×group×time interactions were not significant (Table 4). Fisher’s LSD post-hoc tests showed a significant decrease in the DASS total score only in patients with MS who received active tDCS after the intervention compared to pre-intervention (*t*=2.46, *p*=0.020) but not in the sham group. Compared to the sham group, the active tDCS group showed significantly reduced psychological distress (*t_depression_* =2.73, *p*=0.008, *t_anxiety_* =1.99, df=30, *p*=0.050, *t_stress_* =2.43, *p*=0.017) after the intervention. Baseline between-group comparisons (active groups vs sham) showed no significant differences in the pre-intervention scores (*p*>0.05) (Fig. 3 M-P).

### 3.3. Efficacy of tDCS on cognitive functions in MS patients

#### 3.3.1. Psychomotor speed

Psychomotor speed was evaluated with the RTI task. A significant main effect of time (*F_1,35_*=127.32, *p*<0.001, *η*p2=0.784) and a significant group×time interaction (*F_1,35_*=73.491, *p*<0.001, *η*p2=0.677), but no main effect of group was found for the 5-choice movement time. A similar significant main effect of time (*F_1,35_*=11.684, *p*=0.002, *η*p2=0.25) and a significant group×time interaction (*F_1,35_*=34.992, *p*<0.001, *η*p2=0.50), but a non-significant main effect of group was found for the 5-choice reaction time (Table 4). Fisher’s LSD post-hoc tests revealed a significantly faster movement time in patients who received active tDCS after the intervention compared to pre-intervention (*t* =5.56, *p*<0.001). Additionally, in the post-intervention condition, movement time was significantly shorter in the active group compared to the sham tDCS group (*t* =3.86, *p*<0.001). For reaction time, neither group responded faster after intervention as compared to the pre-intervention condition. However, performance after intervention was significantly faster in the active vs sham group (*t* =2.08, *p*=0.043). Baseline between-group comparisons were not significant in both outcome variables (*p*>0.05) (Fig. 4 A-H).

**Figure 4.**
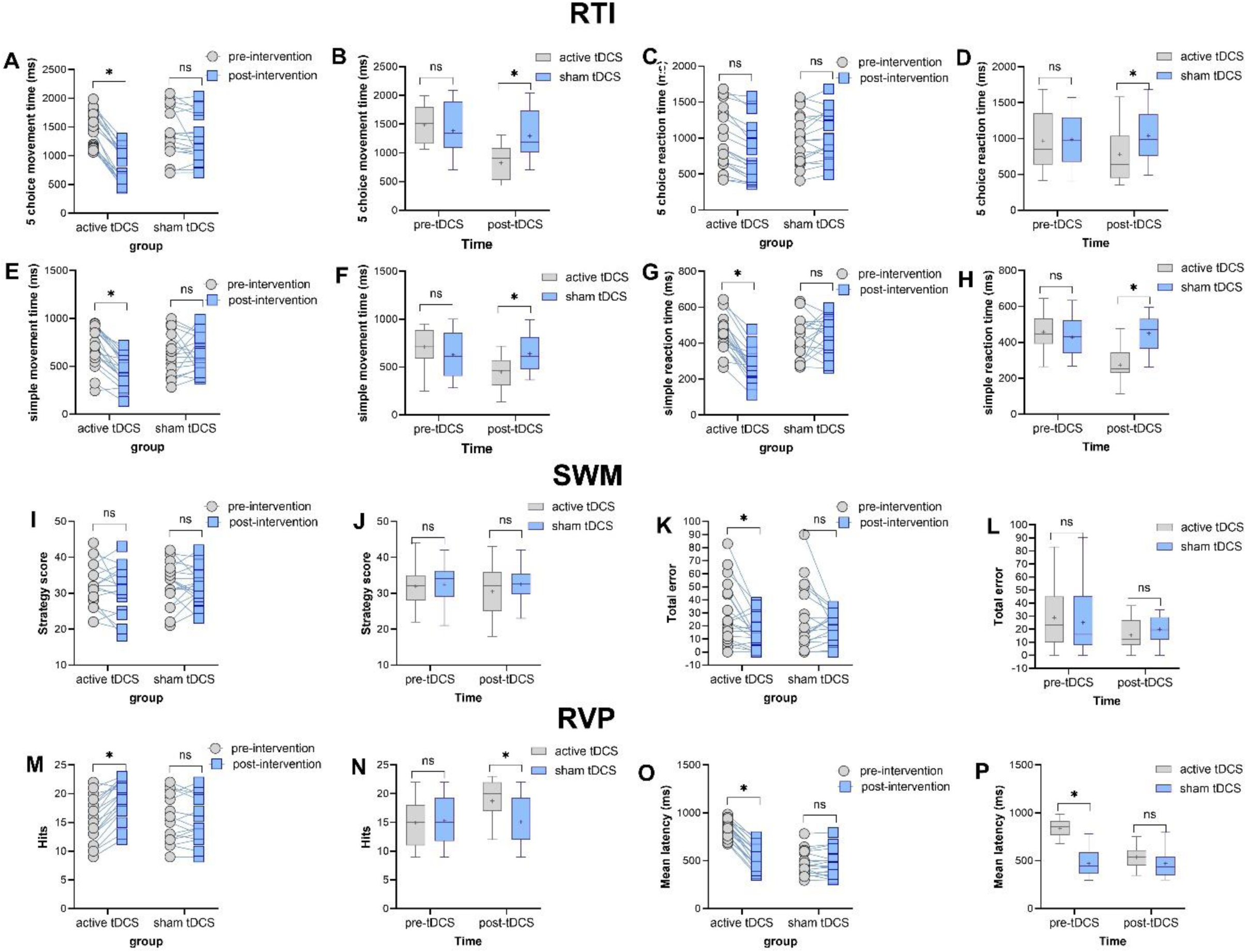
The effect of the intervention on cognition. The left panel displays average outcome measures before and after the intervention within each group (i.e., within-group comparisons). The right panels illustrate average outcome measures for each time point (i.e., pre-intervention or post-intervention) across groups (active vs sham). *Note*: tDCS = transcranial Direct Current Stimulation; RTI = Reaction Time; SWM = Spatial Working Memory; RVP = Rapid Visual Processing; Floating asterisks [*] in the left panel represent a significant difference between pre-intervention measurements vs post-intervention measurements in all groups. Floating asterisks [*] in the right panel indicate a significant difference between active stimulation (1.5 mA) vs sham tDCS at each time point. ns non-significant. All pairwise comparisons were conducted using Fisher’s LSD multiple comparisons tests. All error bars represent s.e.m.

Other outcome variables of interest in the RTI task were *simple*-choice movement time and reaction time. Here, the results of the mixed model ANOVA revealed a significant main effect of time (*F_1,35_*=17.476, *p*<0.001, *η*p2=0.333) and a significant group×time interaction (*F_1,35_*=20.905, *p*<0.001, *η*p2=0.374) but no main effect of group on movement time, and significant main effects of time (*F_1,35_*=24.729, *p*<0.001, *η*p2=0.414) and group (*F_1,35_*=6.439, *p*=0.016, *η*p2=0.155) and a significant interaction between both variables (*F_1,35_*=40.317, *p*<0.001, *η*p2=0.535) with respect to reaction time. Fisher’s LSD post-hoc tests showed significantly faster movement and reaction times in the psychomotor speed task in patients who received active tDCS after the intervention compared to pre-intervention (*t* =4.09, *p*<0.001; *t* =5.60, *p*<0.001). Both movement and reaction times in the psychomotor speed task after intervention were significantly faster in the active tDCS compared to the sham group (*t* =2.92, *p*=0.004; *t* =5.31, *p*<0.001). Baseline between-group comparisons were not significant for both outcome variables (*p*>0.05) (Fig. 4 A-H).

#### 3.3.2. Working memory

Here strategy scores and total errors were analyzed. For the strategy scores, the results of the mixed model ANOVA showed no significant main effects of time, group, or their interaction (Table 4). For the total errors, however, a significant main effect of time (*F_1,35_*=9.081, *p*=0.005, *η*p2=0.206) but no main effect of group or the group×time interaction was observed. Accordingly, no post hoc tests were conducted for errors (Fig. 4- I-L).

#### 3.3.3. Attention and vigilance

The results of the mixed model ANOVA showed a significant main effect of time (*F_1,35_*=39.44, *p*<0.001, *η*p2=0.53) and a significant group×time interaction (*F_1,35_*=47.032, *p*<0.001, *η*p2=0.573) but no main effect of group (*F_1,35_*=1.698, *p*=0.201) on Hits (accurate responses). Patients who received active tDCS had a significantly higher number of hits after intervention vs pre intervention (*t* =2.96, *p*=0.004), and the number of hits was also significantly higher in the real intervention than the number of hits in those who received sham tDCS after intervention (*t* =2.79, *p*=0.006). Similarly, significant main effects of time (*F_1,35_*=291.414, *p*<0.001, *η*p2=0.893), group (*F_1,35_*=32.178, *p*<0.001, *η*p2=0.479) and a significant group×time interaction (*F_1,35_*=292.129, *p*<0.001, *η*p2=0.893) emerged for the mean reaction time. Fisher’s LSD post-hoc test showed a faster reaction time after the intervention as compared to the pre-intervention only in the active tDCS group (*t* =7.28, *p*<0.001). Here, however, mean reaction time at baseline was significantly different across groups, but not after intervention (Fig. 4 M-P).

### 3.4. Improved mental health is associated with cognitive task performance after intervention

We calculated Pearson’s correlations to assess if changes in mental health outcomes are linked to cognitive task performance after intervention and found a significant correlation among specific variables. Notably, enhanced quality of life was significantly associated with faster performance in the psychomotor task (*r*_5choice-move-time_=-0.383, *p*<0.05, *r*_5choice-reaction-time_=-0.481, *p*<0.01, *r*_simple-move-time_=-0.587, *p*<0.01). Improved attention/vigilance (i.e., higher performance accuracy) also showed significant negative correlations with insomnia (*r*=-0.423, *p*<0.05), hypersomnia (*r*=-0.478, *p*<0.01), and overall sleep difficulties (*r*=-0.434) (i.e., higher accuracy was associated with lower sleep disturbances). Additional significant pertinent correlations (e.g., faster psychomotor speed and lower sleep issues) are detailed in Table S1 (supplementary materials).

## 4. Discussion

As a neuroinflammatory disease, MS impairs neuronal efficacy and neuroplasticity. Non-pharmacological approaches, such as non-invasive brain stimulation, have the potential to enhance the outcomes of pharmacological and physical interventions in MS by enhancing neuroplasticity and functional connectivity (42–44) and potentially have anti-inflammatory effects as observed in several noninvasive brain stimulation studies (45, 46), although the latter is not completely clarified. In this randomized, double-blind study, we investigated the effectiveness of repeated prefrontal tDCS in enhancing mental health and cognitive functions in MS patients to assess their impact and potential associations between improvements. Patients who received real tDCS reported a significantly higher quality of life and reduced sleep difficulties after the intervention. They showed also an overall lower level of psychological distress after the intervention compared to the sham group. In addition to these mental health-related variables, patients who received prefrontal tDCS showed superior performance in a neuropsychological test battery that measured psychomotor speed, working memory, and visual attention and vigilance which are frequently impaired in MS patients.

The use of tDCS to improve motor and/or cognitive deficits in MS has been less common compared to other neurological/psychiatric conditions. Moreover, results have been mixed so far. While some studies showed promising benefits in ameliorating fatigue, pain, and cognitive symptoms, but inconsistent effects of tDCS on motor symptoms (23, 24), others show a more promising effect on motor functions (25) which could partially be due to heterogeneous protocols applied in these studies, but also different patient characteristics. This is the first study that specifically investigated the efficacy of tDCS on quality of life, mental health-related variables and cognitive functioning of patients with MS. Our study provides supportive evidence for the efficacy of tDCS in improving quality of life and amelioration of psychological and cognitive deficits in patients with MS. Here, the applied protocol and proposed mechanisms of effects are discussed.

Prefrontal tDCS has been extensively used to improve cognitive and emotional impairments in clinical populations, as well as to enhance cognitive abilities in healthy individuals. Prefrontal tDCS can have pro-cognitive effects in neuropsychiatric disorders that are characterized by executive and cognitive deficits, such as mood and anxiety disorders (47, 48), obsessive-compulsive disorder (49, 50), and substance use disorder (51, 52). Pro-cognitive effects were also shown in neurological disorders such as Parkinson’s disease and stroke (53, 54) and major neurodevelopmental disorders such as ADHD and autism (55–57). It has been also effectively used for improving fatigue and quality of life (58, 59). The rationale behind the use of prefrontal tDCS for enhancing cognition mostly comes from the involvement of different regions of the prefrontal cortex in various aspects of cognitive functions (60–62) which seems to be at least partially applicable for MS.

Cognitive impairment in MS is the consequence of widespread lesions in the brain and frequently includes deficits in complex attention, efficiency of information processing, executive functioning, processing speed, and long-term memory (4, 16). Recent large-scale neuroimaging studies conducted in MS patients have shown a causal relationship between the pathophysiology of MS and different brain regions, including the frontal and prefrontal cortices (specifically the orbitofrontal cortex) and other regions that are connected to these cognition-related areas (such as the parahippocampal gyrus) (21). Our study applied prefrontal tDCS over prefrontal regions (DLPFC and right FPC) which are important for cognitive deficits in MS, leading to improved psychomotor speed and attention/vigilance in patients who received active stimulation. In addition to the cognitive improvement, quality of life of the patients was improved after the intervention. The frequency of sleep difficulties and states of depression and anxiety were also significantly reduced after intervention. Sleep improvement after the active tDCS can be explained by the more recently stressed role of the cortico-thalamo-cortical feedback loop, which is a top-down regulatory system related to cortical areas (e.g., PFC), in regulating arousal and sleep (63). Mood alleviating effects can be partially explained by enhanced cognitive control as a result of tDCS over the DLPFC, which regulates mood and valence of emotional experiences (64, 65). Importantly, there is a link between the prefrontal cortex, specially the DLPFC, the medial PFC, and the amygdala network in MS patients who experience emotional difficulties (66), and reduced depression, anxiety, and distress following intervention might be explained via this prefrontal-amygdala related emotion regulation. Briefly, Upregulating DLPFC activity can reduce amygdala activity (67), which is usually hyperactive during negative emotional processing in MS (66). This effect, coupled with the functional connectivity-enhancing effect of tDCS (68), may be a possible mechanism of action.

The current study’s findings should also be compared with previous tDCS studies in MS patients. Recent systematic reviews and metanalyses of tDCS studies conducted in MS show inconsistent results about the cognitive effects of tDCS with some studies reporting clear benefits in ameliorating cognitive symptoms (23), some partial improvement in specific aspects of cognition (e.g., vigilance) (25) and some with no strong evidence for effectiveness of tDCS on cognition (24). Previous tDCS cognition-related studies have commonly targeted the left DLPFC with the anodal polarity while using the return electrode over the right DLPFC, right shoulder, or right supraorbital area, and stimulation intensity and duration ranged from 1.5 to 2 mA and from 20 to 30 min respectively (25). The present study aimed to shed light on the effects of tDCS on specific secondary deficits, but also on the interplay between cognitive functions and mental health domains, which are both significantly impacted by MS, but have been less studied in previous works. Importantly, here we observed significant correlations between mental health-related outcome variables and cognitive performance, suggesting that these two impaired domains in MS (mental health and cognitive deficits) are interrelated. One consideration regarding the protocol in the present study (anodal F3-cathodal Fp2) is the use of the reference electrode over the right FPC instead of the right DLPFC. The bilateral DLPFC protocol can also be applied if cognitive functions are the main focus, as indicated in previous tDCS studies in MS (25) and other neuropsychiatric disorders (69, 70). In this study, we also aimed to explore the impact of the intervention on participants’ emotional experiences. FPC, which includes the orbitofrontal cortex (38), was selected over the right DLPFC due to its significant role in emotional regulation and direct connections with subcortical regions (60). The relative efficacy of DLPFC-FPC versus bilateral DLPFC tDCS needs to be compared to determine which is more effective in enhancing mental health and cognition-related variables.

### Limitation

We acknowledge that our study has some limitations. Firstly, although the sample size of this study is larger than the majority of the previous tDCS studies in MS (24, 25), it is still relatively small, and therefore, these findings need to be confirmed in larger trials in the future. Secondly, we did not measure outcome variables over a reasonable follow-up period to determine how long the observed effects lasted and the intervention was not applied daily but every second or third day, which might have limited efficacy. Finally, we did not include any physiological measures, which would have been informative to understand how brain functions and physiology are affected by the intervention and how they align with the behavioural results. These limitations should be considered when interpreting the results of our study and for future larger trials.

## Conclusions

In conclusion, in this study, we upregulated the activity of the left DLPFC and downregulated the activity of the right FPC in 37 patients with MS and evaluated the quality of life, sleep difficulties, psychological distress, and cognitive functions after 10 sessions of brain stimulation. The group that received real intervention showed significant improvement in quality of life, and reduced psychological distress and sleep difficulties, as compared to the placebo group. This improvement was linked to an enhancement of psychomotor speed and attention/vigilance of the patients in the real intervention group. These findings suggest that prefrontal tDCS targeting the left DLPFC and right FPC has the potential to be a valuable intervention for treating secondary symptoms (mental-health-related variables and cognitive deficits) in patients with MS.

## Data Availability

All data produced in the present study are available upon reasonable request to the authors after publication of the peer-reviewed version

## List of abbreviations

MS: Multiple Sclerosis
tDCS: transcranial direct current stimulation
MSIS-29: Multiple Sclerosis Impact Scale
MSQ: Mini Sleep Questionnaire
DASS-21: Depression Anxiety Stress Scale-21
RTI: Reaction Time task
SWM: Spatial Working Memory task
RVP: Rapid Visual Information Processing task
DLPFC: dorsolateral prefrontal cortex
FPC: frontopolar cortex

## Declarations

## Ethics approval and consent to participate

All patients gave their written consent to participate in the study. The protocol was conducted in accordance with the latest version of the Declaration of Helsinki and was approved by the Institutional Review Board and Ethical Committee at Mohaghegh Ardabili University.

## Consent for publication

Not applicable

## Availability of data and materials

The datasets used and/or analyzed during the current study are available from the corresponding author upon reasonable request.

## Authors’ contributions

NZ: conceptualization, investigation, data curation, visualization, validation; SB & HGL & MN: supervision, resources, project administration, data curation; MAN: writing—review & editing; MAS: supervision, methodology, writing original draft, writing—review & editing, visualization, formal analysis. All authors read and approved the final manuscript.

## Acknowledgments

We would like to thank the MS Association located in the city of Rasht, Iran for their assistance in recruiting patients.

